# COVID-19 and Medical Education in Ghana: Assessing the Impact

**DOI:** 10.1101/2021.03.11.21253306

**Authors:** Edward Asumanu, Linda Tsevi

## Abstract

Medical education in Ghana has been affected in many ways by the onslaught of the COVID-19 pandemic. Though the pandemic has affected both preclinical and clinical segments of medical education, the effect has been felt more at the clinical stage. Medical students on vacation who started their clinical training abroad could not return to their destination of study to complete their programme because of COVID-19 linked travel restrictions. This qualitative study examined how COVID-19 impacted on teaching and learning at a public higher education institution offering clinical medical education in Ghana for over 200 medical students. These medical students were from three different higher education institutions with varied curriculum outcomes. Thus, for them to be considered as a single group required innovativeness on the part of administrators. Open-ended interviews were held with administrators and the outcome indicated that salient aspects of the clinical training process had been impacted. These included administration of clinical education, curriculum, student learning, student assessment and code of practice. As a result of the pandemic, student learning shifted from traditional face to face interaction to online learning at the beginning. Some of the administrative challenges that ensued included the need for reduced number of students per tutor and introduction of afternoon sessions with a limited budget. The paper concludes that COVID-19 has been disruptive to traditional medical education in Ghana. However, the novel learning processes may provide opportunities to increase access to medical education using a phased system of learning. The findings from this study should have implications for policy and contribute to the discourse on blended learning in medical education in Ghana while ensuring that quality is maintained in all instances.

## Introduction

COVID-19 pandemic has precipitated a number of transformations in the education sector globally and the medical education unit has not been spared [1]. Clinical and practical components of medical education that had been handled on a face-to-face basis for decades had to be suddenly revised and transformed into a blend of online and limited face-to-face programmes while ensuring that COVID-19 protocols were observed to guarantee faculty and student safety. Suffice it to say that in the Sub-Saharan African country of Ghana, the global COVID-19 pandemic has disrupted the traditional model of medical education which, over a long period of time has been very conservative in its structure and function [2].

Overall, the COVID-19 pandemic has caused an overhaul of medical school orthodoxy that whilst disruptive, may serve to expose institutions to novel means of teaching and assessment which may ultimately improve medical education in the future [3]. In Ghana, COVID-19 has interrupted the clinical aspect of medical education in many ways. During a three-week lockdown in Ghana which ended on April 19, 2020 clinical medical education was suspended just like any other educational activity. After the lifting of the three-week lockdown and easing of some restrictions, medical students in higher education institutions undertaking clinical components, transitioned to a blended form of learning. Initially, medical students were reluctant to attend classes for fear of contracting the virus even with all the safety protocols in place, and this affected teaching and learning. However, with counselling and assurance from administrators of these institutions, the students gradually eased into the learning mode. The students also attended classes with new codes of dressing proffered by the administration. Findley [4] notes that when students returned to clinical rotation eventually, all didactic teaching have evolved to online formats. Moreover, clinical experiences of students had to come with the wearing of personal protective equipment and maintenance of social distance. Faculty members and administrators also had to be creative in their clinical supervision of students.

In line with safety protocols, there were fewer number of cases for teaching the clinical component in Ghana’s higher education institutions. In addition to merging some of the lessons, administrators ensured that class sizes were reduced to prevent infection while e-learning was rapidly deployed as a means of teaching. Research indicate that the beginning of the pandemic led to a suspension of didactic teaching to be replaced by live-stream synchronous or recorded lectures which negatively impacted collaborative experiences and communication skills of students [5,4,6].

Medical education in Ghana is a five and a half year programme consisting of preclinical and clinical stages. The clinical stage of training includes in-hospital clinical rotations, theoretical lectures and at the same time revolves around patient interaction which aids in understanding the pathology of diseases. Traditionally it was structured on face-to-face communication (history taking) and physical contact (physical examination). This method of teaching thus required close contact with patients whose diseases were not predetermined. Indeed the essence of the clinical training stage is for the student to interact, determine the disease and plan an appropriate management. Since COVID-19 has been known to have a direct human to human transmission as well as a high morbidity and mortality rate the traditional clinical training mode was affected. One key mode of transmission is contact with an infected person in an unprotected manner. However, as most infected persons may be asymptomatic, preventive measures required distancing and reduced contact. The combination of the disease transmission, its fatality and preventive measures have greatly impacted the clinical stage of medical education. According to Findley [4] classes for medical students in clinical training in Texas, U.S.A. who were inured to the hands-on nature of clinical medical education, were suspended to among others, protect students and ensure social distance in clinical settings. Notably, the transition to a blend of both online and limited face-to-face contact was not without its challenges as students needed this fusion to enable the efficient practical application of theories learnt [5, 4, 6].

Invariably, the COVID-19 pandemic has obliged administrators of higher education institutions to be innovative in their ability to replicate some of the face-to-face clinical contact and engagement into a virtual environment. Thus, this paper aims to describe and explain the experiences of a higher education institution offering clinical medical education during the COVID-19 pandemic in Ghana. These experiences have come with challenges and opportunities for administrators, faculty members and students. It is worthy of note that a number of publications have examined student experiences in this pandemic (5, 4, 7] but there is a dearth of papers on how institutions offering medical education have coped with the pandemic [8] especially in the Sub-Saharan African region generally and Ghana in particular. This paper therefore attempts to bridge this gap by offering a Ghanaian perspective about innovative practices adapted in teaching and learning in view of resource challenges that higher educational institutions in developing countries are encountering in the midst of the pandemic. Three research questions guided this study: (a) How has departmental administrators of a higher education institution offering clinical medical education in Ghana adapted teaching and learning to the COVID-19 pandemic? (b) How did the administrators overcome challenges associated with the shift to a blend of online learning and limited face to face interaction? (c) How were the clinical rotations of three different groups of students managed simultaneously?

## Method

This qualitative study purposefully selected and interviewed six departmental administrators from a Ghanaian public higher education institution that has provided clinical training to undergraduate medical students for a number of decades. The study employed an in-depth unstructured open-ended interview protocol. The objective of the interview method was to obtain salient information from the administrators regarding the processes that had gone into the provision of blended learning to medical students undergoing clinical training in the era of COVID-19. Interviewing is an effective means to gain insights into experiences and perspectives of administrators in a higher education institution [9]. Similarly, according to Rubin and Rubin [10] interviews are very beneficial because they enable the researcher to decipher what has happened, why and what it largely means. The 37 Military hospital was selected for the study because it is a tertiary facility that provides final year clinical training for undergraduate medical students in surgery, obstetrics and gynaecology, paediatrics and internal medicine trained both in and outside the country. It is significant to note that during the COVID-19 pandemic in 2020, the hospital had over 200 students from three higher education institutions (namely a public, private and a foreign medical school) undertaking clinical rotations simultaneously and at various stages of training. Students from the public and private medical schools were taking their regular clinical courses while the foreign medical students were included due to air travel restrictions during the pandemic. Previously these rotations were staggered such that each category had its own period of clinical rotation. The 37 Military hospital coordinated with all three institutions and adapted its own situation to facilitate the provision of clinical training programmes. The clinical programme is supervised by the undergraduate unit of the hospital and implemented by six clinical departments namely Anaesthesia and Intensive Care Units, Obstetrics and Gynaecology, Medicine, Paediatrics, Public Health and Surgery. The Department of Surgery is made up six core units namely Cardiothoracic, General Surgery, Orthopedics and Truama, Plastic and Reconstructive, Urology and Neurosurgery. Other associated units are Opthalmolgy, Ortholaryngology and Maxillofascial. The Medical Department has subspecialty units including Cardiology, Nephrology, Diabetes and Endocrinology, Respiratory, Nephrology, Dermatology, Psychology, General Medicine and the Haematology unit. The Paediatric unit has a Neonatal Intensive Care unit and a Paediatric emergency unit. It runs specialist clinics in Cardiovascular, Sickle Cell, and General Paediatrics. The Obstetrics and Gynaecology Department has separate Gynaecological and Obstetrics emergency units. The Public Health Department has all the components of preventive health and has clinics and programmes from mother and child welfare as well as infectious disease clinics. This unit is involved in community health and public health education campaigns

In this study, all six participants were recruited by the lead researcher in October 2020 from the departments that constitute the undergraduate unit. Telephone and face-to-face interviews as the situation demanded were conducted while obeying all the COVID-19 safety protocols. These contacts with participants were made after the clinical training sessions for final year students had ended. The study was also approved by the lead researcher’s Institutional Review Board (IRB) with reference number 37MH-IRB/NF/IPN/467/2020. An unstructured open-ended interview protocol served as a guide for the data collection. The lead researcher conducted all interviews in person and permission was sought from the participants to audio record. When interviews are recorded, it offers an accurate data capture [11]. In order to maintain confidentiality, actual participant names were not included in the study. The duration of each interview ranged from 50 minutes to about an hour and a half. Copies of the interview protocols, which were informed by the research questions guiding the study, were given to the participants prior to the interview date. Essentially, the interview schedule for the administrators focused on how they have adapted teaching and learning to the era of COVID-19 and addressed challenges encountered among others.

The transcripts from the interviews with each participant were coded separately through the application of content analysis by using both inductive and deductive approaches [12]. The coding was done manually by reading through the interview transcripts several times to look for repeating ideas that were relevant to the research questions among others. Each interview transcript was read at least three times to acquaint the researchers with the themes that would emerge from the coding process. The researchers also undertook open coding of the transcripts to be followed by axial coding by developing categories. The researchers examined themes that emerged from the interviews both deductively and inductively and looked for linkages and patterns among themes. Descriptive categories were identified and copies of the interview transcripts and emergent themes were sent to participants by email. The participants acknowledged that the interpretation of the data was accurate after reviewing the transcripts. To further ensure reliability and validity of data analysed, there was member checking thus, a peer reviewer reviewed all the codes to eliminate possible research bias.

## Results

The outcomes of the study are reported under the following thematic areas namely: administration of clinical education, student learning, curriculum, student assessment, code of practice, opportunities and challenges

### Administration of Clinical Education

The participants noted that they had to think outside the box to enable the smooth running of the clinical component of education as well as address some of the challenges that this pandemic has brought about. The clinical part of medical education is indicated as a very critical component of the training process, without which a medical student will not be able to graduate and practice. A participant from the undergraduate unit noted that discussions were held with stakeholder establishments regarding the clinical rotation of medical students and the need for those in their final year to complete their rotations and graduate. However, there were challenges associated with the attainment of this objective. One participant that we will call Kodjo stated:

> The common theme among all the schools was the need to complete rotations and graduate their final year group. The challenge for our institution was the aggregate large number of students from the beneficiary institutions….. The 37 Military hospital had adequate infrastructure and technology backbone to mitigate the impact of the COVID-19 but there were still challenges with inter and intradepartmental implementation of programmes. A centralised planning was done for the overall implementation of programmes.

For very valid reasons, participants indicated that they had to find ways of balancing the need to complete the academic calendar despite the substantial time lost because of the pandemic, while incorporating social distance into the programmes. Student groups had to be reduced from an average of 10 per tutor to five for clerkship rotation. Afternoon sessions which were not the norm, were introduced to accommodate the increased student groups created as a result of the pandemic. Another administrative concern that had to be addressed was recruiting tutors on short term basis to meet the demands of COVID-19 protocols. This had to be undertaken without an increase in either departmental or institutional administrative budget. Some of the innovative activities that participants had to implement include ensuring reduced face-to-face interactions and long hours of screen time [13]. Majority of the participants agreed that the success of this blended form of clinical education may continue to be the new normal for a while. However, it was imperative that the preliminary challenges encountered were addressed efficaciously so that medical students get the best out of this experience and eventually contribute their quota to the health field in the long run. Though there is no clarity on the overall impact of COVID-19 on medical education and how the current adaptions may influence its future, participants were of the view that the innovative ways had yielded positive results [14]. Kwaku, a participant stated: *Staggering the different groups’ clinical rotation times was a useful approach to managing the large number of students while adhering to the COVID-19 safety protocols*.

### Student Learning

Student learning has changed drastically in the era of COVID-19. As earlier indicated the common mode of learning in clinical years in Ghana is by clerkship rotation at various clinical wards, outpatient departments and theatres. The participants noted a need for modification to two components of clinical teaching consisting of practical sessions and classroom lectures/tutorials. Practical sessions were modified to have reduced numbers while aspects of the classroom sessions were changed from face to face to online teaching. However, the student numbers for the online teaching were kept unchanged. Before COVID-19 struck, faculty members had to contend with large class sizes in didactic lectures and clinical rotations. There was also institutional adoption of the creative use of video conferencing, social media platforms, and usage of free open access medical tools. These were used to maximum effect at departmental levels with Zoom video conferencing being the most common method adopted [15, 16] according to majority of participants. Effecting technology into medical education in Ghana, though distinct, had its challenges. Some of the challenges encountered by students included bandwidth issues and reliable and stable internet connectivity [16]. On a significant note, participants also observed that student attendance and utilisation of library facilities increased during this COVID era.

Kwame (not his real name), a participant, made an important observation about faculty members’ usage of technology. He stated: *Younger faculty members found online learning exciting and quickly adapted. On the other hand, older faculty members had initial difficulties adapting*. Further, there were limited clinical experiences sessions for students mainly because of the imposed nationwide restrictions. These restrictions required medical educators to outline priorities for the limited clinical experiences and design different approaches to competency attainment [17]. Kwame further noted that: *During the pre-COVID era, practical sessions for medical students were held mainly in the morning but with the onset of the pandemic, afternoon clinical training sessions were added to cater for the increased numbers in student groupings*.

The COVID**-**19 protocols that had to be adhered to included social distancing rules and the wearing of personal protective equipment (PPE). Since the disease can cause life threatening conditions instructors must deliver lectures safely, while ensuring the integrity and continuity of the medical education process [18]. Consequently, participants were of the view that emergency units that used to be a critical site for training, should be excluded because of COVID-19, and this limited the number of cases for learning. They also note that before the pandemic, students were expected to have daily contacts with patients but this was reduced and replaced by tutor-led demonstration of clinical signs of diseases. During these demonstrations, the tutors directed students on what to do and what to find on a patient. In the pre-COVID era, students were to elicit their own findings based on lessons taught in the classroom that encouraged a more hands-on process while the tutor corrects mistakes. The present system under COVID, uses observation of patients by students to acquire the skill needed for practice which is not desirable. It has been reported that in teaching by observation, clinical learners are passive observers which indicates a low yield use of an invaluable resource [19]. As an alternative to hands-on procedures, the institution introduced the use of dummies and mannequins as replacement for live patients. During the clinical training process, medical institutions in Ghana needed to ensure that students are exposed to a wide range of cases including those that present as emergencies. But COVID-19 changed this traditional learning trajectory as exposure to all emergencies was drastically reduced. Thus medical students did not encounter many emergencies such as heart failure and ectopic pregnancies. Moreover, administrators of medical institutions in Ghana had to ensure that switching partly to online learning would not adversely affect student learning outcomes.

### Curriculum

Data from the study indicates that the curriculum in clinical medical education in Ghana was modified because of COVID-19. Sani [8] in examining the consequences of the pandemic on curriculum delivery and student assessment in the UK, noted that many of the written assessments were changed to open-book examinations by medical schools in response to the COVID-19 pandemic. This raised a possible issue of examination being an inadequate measure of students’ theoretical knowledge due to ease of access to online resources. Overwhelmingly, the participants believed that as a result of COVID-19, curricula for the clinical stages had to be redesigned to highlight the essential requirements that meets the minimum standards of training. Changes had to be made to both content and duration components of the course structure. It was a challenging task striking a balance between the different curriculum requirements for the three groups of students. A participant that we will call Ama noted:

> The institutions made changes to curriculum used for training medical students. At our Department, the mode of delivery of the curriculum content varied and changed from online to demonstration teaching. The different institutions had varied curriculum for the same clinical rotation.

In most instances about a quarter of the proposed programme had to be omitted according to majority of the participants. This was to make up for time lost in the academic calendar. As part of the innovations, lectures were done online at the beginning of the clinical rotation and the practical sessions followed. This was intended to keep students away from the patients as much as possible. Previously, lectures and practical sessions were held concurrently but with this innovation, contact between students as well as the risk element was reduced by half. It is anticipated that ingenuities of medical education will emerge out of the COVID-19 pandemic [20]. Ultimately, curriculum revision resulted in reduced duration of contact and content. Research further indicates that the beginning the pandemic led to suspension of didactic teaching to be replaced by live-streams or recorded lectures. And this negatively impacted collaborative experiences and communication skills of students [5, 4, 6].

### Student Assessment

The goal of student assessment during clinical years is to meet requirements of the Medical and Dental Council in Ghana and the universities of training. Prior to the onset of the pandemic medical students’ assessment in Ghana was gradually shifting from the traditional long case and short case type of assessment to the Objective Structured Clinical Examination (OSCE). Notably, the participants were of the view that pandemic has accelerated the introduction of OSCE into student assessment in medical education in Ghana. While OSCE was suitable for the pandemic, a participant (Ama) stated:*…*.*it had its drawback in difficulty with implementation and maintenance of high standards set by most medical schools*.

As a result of COVID-19, practical assessments had to be rescheduled; the implication being that students had longer periods to prepare adequately for the practical component. The participants further noted that clinical students’ end of rotation assessment took the form of multiple choice questions and short answer clinical-based questions. Practical clinical examinations and oral interviews were excluded.

### Code of Practice

Data from this study indicates that the advent of COVID-19 has brought swift changes in the code of practice for medical education in Ghana. The code of practice comprises student dressing and preparation for clinical sessions among others. Participants noted that they had to be creative in their supervision of students both theoretically and during practical sessions. Previously, the dress code of medical students had traditionally been orthodox and conservative where the student is expected to be attired in a formal dressing with a white overcoat. But with the advent of the pandemic it meant that students have to appear in “scrubs” (short sleeve dresses used for surgical operations) to protect them from infection. In order for stakeholders to accept the current reality of new dress modes, they had to be formally informed. One participant called Abena intimated that: *It needed internal communication to have all stakeholders accept the current reality*. The reason being that this was an alien practice to faculty members, hospital staff and clients attending hospitals. It thus portrayed a sociocultural phenomenon of how society perceived medical students in training as well as doctors.

It is significant to note that the use of the open book method of learning in wards and clinical settings was not part of the traditional way of teaching and learning in Ghana. Students were expected to have undertaken their reading before an outpatient ward or theatre session and apply this knowledge acquired to practical scenarios but the pandemic changed this tradition. COVID-19 also provided students with the opportunity to use technology to find information during practical sessions. While the use of technology made some sessions disruptive because students were online searching for answers, other tutors used the opportunity to motivate students to research and stimulate self-directed learning at practical sessions. But some of the participants indicated that students had difficulties with older faculty in this instance because they regarded the use of gadgets in clinical sessions as inappropriate behaviour. There was also a challenge regarding patients voluntarily offering themselves to be used for practical sessions during this COVID-19 period. A participant that we will call Kofi stated: *Patients were not used to medical students searching for information online during clinical training sessions. This affected their willingness to volunteer for teaching purposes*.

Majority of the participants noted that privacy, a key component of medical interaction, became a challenge due to fewer number of available patients. Therefore, students had to be paired and patients had to be reused multiple times for training sessions. Providing medical and personal history to more than one student for multiple times compromised privacy of patients. The situation worsened when patients have to be subjected to physical examination in the same manner. Eventually, the consequences were that parts of the examination were excluded from the teaching sessions. A participant (called Kodjo) noted: *It is yet to be determined the effect of these practical sessions on the ability of trainees to perform these examinations and make interpretations appropriately*.

The elimination of some aspects of clinical education as a consequence of COVID-19 led to a feeling of vulnerability among students as they could not be of immediate assistance to their faculty members in the discharge of their clinical duties. The students did not partake in frontline service provision perhaps from the fear of contracting COVID-19. However, the students found ingenious ways of assisting medical personnel and were not directly involved in the frontline service provision. In Singapore, medical students were involved in the design of workflows for patient management [7]. Findley [4] also indicates that when students returned to clinical rotation eventually, all didactic teaching have evolved to online formats. Students had to wear personal protective equipment while maintaining social distance and obeying all the safety protocols.

### Opportunities and Challenges

The COVID-19 pandemic has brought about opportunities as well as challenges in clinical medical education in Ghana. Research indicates that the establishment of small groups or interactive learning communities through leverages in technology such as webinars and videoconferences has enhanced student learning experiences in this era while quality is not sacrificed [5, 7, 8]. This was largely the observation in this study. Another major impact that COVID-19 had on medical education is the cancellation of face-to-face conferences which has affected medical students who require it in their applications for residency. This has resulted in the demand for online conferences to enable prospective students participate. The pandemic has therefore provided a need for leveraging technology such as videoconferencing, teleconferencing, webinars and the usage of e-learning platforms for both undergraduate and postgraduate medical education [5, 4, 6].

Nonetheless, Jaideep [6] is of the view that though teaching online during the pandemic is now the new normal, it comes with challenges such as students’ inability to sufficiently meet their learning goals in relation to internal assessment examinations and clinical ward rounds among others. A participant indicated that some sessions were merged and others were not completed as a result of the pandemic. This was done to enable final year medical students to graduate. The suspension of in-person didactic class sessions among others, negatively impacted on the capability of students to develop collaborative, clinical and communication skill sets as well as group learning [5, 4, 7, 6]. Equally, a number of noticeable challenges surfaced during this period. Majority of the participants indicated that as a result of multiple student sessions at the clinical stage, there was the need for increased numbers of faculty members and support staff though funding was not readily available. Also, it was sometimes difficult to track student attendance online and attitude to practical sessions as there was noticeable fatigue among faculty members resulting in cancelled lectures and practical sessions. There was additional financial strain on the departments as administrative staff who shared a common office had to relocate to a new building so that social distancing protocols could be followed to prevent the spread of COVID-19. This required the use of additional resources and finances that had not been budgeted for. Financial constraints and work output of support staff continue to be some of the pressing challenges in managing medical education [21]. Kwame, a participant stated:….. *One other noticeable issue was that the work output of administrative staff working from home in alternate weekly fashion decreased when compared to work output in the pre-COVID era*.

## Discussion

This study examined the impact of COVID-19 on clinical medical education in Ghana from the perspective of administrators of a higher education institution in Ghana, a country in Sub-Saharan Africa. The outcome of the study noted themes such as administration of clinical education, student learning, curriculum, student assessment, code of practice, opportunities and challenges in the assessment of COVID-19’s impact on medical education in Ghana. Some of the perspectives noted in this study aligned with what pertains globally [22]. The participants generally noted that they had to think outside the box regarding clinical rotations as well as determining innovative ways of completing the academic calendar to enable medical students graduate and complement the national health workforce. The participants for the first time, had to tussle with three student groups of varied backgrounds concurently. Thus they had to run three different curricula simultaneously in a midst of a global pandemic. The study further indicated that student learning changed from a face to face didactic component to a blend with online and expansive use of technology. Younger faculty members seamlessly adapted to technology usage as opposed to older faculty members who had issues during the transitioning process. Students’ challenges with technology usage had to do more with bandwidth issues, reliable and stable internet for online classes rather than skill. Participants further noted that the regular class sizes also had to be reduced to ensure that COVID-19 safety protocols such as social distancing and wearing of personal protective equipment were adhered to. The reduction in class sizes however, came with faculty fatigue as they had to do more sessions than they were previously accustomed to.

Emergency cases were also reduced in clinical training and this impacted on the collaborative learning skills of students [5, 4, 6]. The adaption in practice where students have to appear in protective equipment and scrubs aggregating around a patient was new to our sociocultural practices. The absence of emergency cases for clinical education needs to be studied further to determine its impact on performance in practice. This study highlights the need for a harmonised curriculum among medical schools for effective administration of course content during clinical years. The content should meet the minimum standard of care while preparing students for new and unanticipated global emergency.

Student assessment also shifted from the traditional long case assessment to the Objective Structured Clinical Examination (OSCE). This was a rapid paradigm shift that students, faculty members and administrators adapted to. The data suggests that though administrators were ill prepared in terms of resources and training to undertake the OSCE but were able to achieve the desired objective. OSCE is an expensive method for clinical assessment and the cost must be adjusted in medical education funding. The use of OSCE as an assessment tool is suitable in times of emergencies and provides a practical method in the absence of patients. Findings from the study also indicate opportunities and challenges that participants had to address. Some of the opportunities included the leveraging of technology for videoconferencing, webinars and e-learning platforms for medical education as a result of the pandemic [5, 4, 6]. Technology also had to be leveraged by the institution to foster synchronous teaching and a limited face-to-face component, though it did not come without its challenges such as quality of internet streaming [5, 4, 6, 23]. The code of practice relating to student dressing changed as a result of COVID-19. The surge in technology usage in clinical medical education has become the new normal. There is now a rise in online conferences, interactive learning communities, webinars and video-conferences among others through leverages in technology usage. These have enhanced student learning experiences in the era of the pandemic while quality is not totally sacrificed [5, 7, 8]. There is an anticipated prolonged lifespan of physical infrastructure from reduced usage and subsequent wear and tear.

Challenges that participants indicated included inability to track student attendance since some class sessions were held online. There was also limited opportunity to evaluate attitude to practical sessions. Other challenges were inadequate funding and resources, and faculty fatigue emanating from numerous class sessions scheduled to make up for lost time. The participants noted that they had to find ways of balancing the need to complete the academic calendar despite time gone, while incorporating all the needed safety protocols into the clinical programmes. There were reported long hours of screen time and reduced face-to-face interactions [13, 14]. Primarily before the onset of the pandemic, medical education in Ghana had traditionally been a structured face-to-face communication. However, this structure has changed in the COVID-19 era as administrators had to be innovative to continue the clinical component of medical education. Moreover, the clinical experiences of students had to come with the wearing of personal protective equipment and maintenance of social distance [14]. Some older faculty members were not too happy with the opportunity given to students to look for answers online during clinical sessions since they felt it was interrupting its smooth flow.

### Limitations of Study

This study has several limitations in that the participants were purposefully selected from one public institution providing clinical training for medical students from three higher education institutions. Thus the findings may not be entirely generalizable. Second the number of participants was small and the study only focused on experiences and perspectives from administrators. Future studies could investigate student and faculty member experiences and perspectives from both public and private institutions. The experiences of student and faculty member could be compared with that of administrators’ to determine if there were any similarities and differences. These findings could also aid institutions offering clinical medical education in Sub-Saharan Africa as to what will work best in Africa’s peculiar environment.

## Conclusion

This paper has examined innovative ways adopted by a public higher education institution in Ghana offering clinical medical education to three higher education institutions during the COVID-19 pandemic using a blended format. A number of innovations had to be implemented to ensure student graduation. Though some of the implemented programmes were not without challenges, the incorporation of technology greatly enhanced the training process and aided in completion and graduation while taking cognizance of student assessment, curriculum and administration among others. As students, faculty members and administrators adapt during this pandemic, technology continues to be leveraged and novel initiatives improved on while quality is not sacrificed. This paper has shown that COVID-19 has been disruptive to traditional medical education in Ghana. However, the innovative learning processes may provide the opportunity to increase access to medical education through the introduction of a phased system of learning. An alternative to foreign medical training will also have to be evaluated if part of training can be undertaken in-country. The findings from this study should have implications for policy and contribute to the discourse on blended learning in medical education in Ghana while ensuring that excellence is maintained in all instances. It is believed that the discussions in this paper may be of significance to other medical institutions in the Sub-Saharan African region.

## Data Availability

The data used to support the findings of this study are included in the article.

## Data Availability

The data used to support the findings of this study are included in the article.

## Conflict of Interest

The authors declare they have no conflict of interest.

## Acknowledgement

The authors acknowledge the invaluable responses from the administrators of undergraduate units of Departments in the 37 Military Hospital during the study period.

## Notes

### Competing Interest Statement

The authors have declared no competing interest.

### Funding Statement

No external funding was received for this study.

### Author Declarations

This study was approved by the 37 Military Hospital Institutional Review Board (IRB) with reference number 37MH-IRB/NF/IPN/467/2020.

